# Aficamten Reduces Eligibility for Septal Reduction Therapy in Obstructive Hypertrophic Cardiomyopathy: Long-Term Outcomes from FOREST-HCM

**DOI:** 10.64898/2026.07.08.26357594

**Authors:** Ahmad Masri, Benjamin Meder, Lubna Choudhury, Pablo Garcia-Pavia, Theodore P. Abraham, Roberto Barriales-Villa, Ozlem Bilen, Perry Elliott, Albert A. Hagege, Sherif F. Nagueh, Srihari S. Naidu, Michael E. Nassif, Iacopo Olivotto, Artur Oreziak, Anjali T. Owens, Omar Wever-Pinzon, Florian Rader, Albree Tower-Rader, Justin Godown, Stephen B. Heitner, Daniel L. Jacoby, Stuart Kupfer, Fady I. Malik, Regina Sohn, Jenny Wei, Sara Saberi, FOREST-HCM Investigators

**Author notes:** **Corresponding author:** Ahmad Masri, MD, MS, Hypertrophic Cardiomyopathy Center, Knight Cardiovascular Institute, Oregon Health & Science University, 3181 SW Sam Jackson Rd, Portland, OR 97239.

## Abstract

**Background:** Septal reduction therapy (SRT) is recommended in drug-refractory, symptomatic obstructive hypertrophic cardiomyopathy (oHCM). We evaluated whether aficamten, a novel cardiac myosin inhibitor, can reliably transition guideline-eligible SRT candidates to ineligibility, and the associated safety profile of aficamten in this group.

**Methods:** We analyzed participants with oHCM enrolled in FOREST-HCM (NCT04848506), the long-term open-label extension study of aficamten, from 28 May 2021 to 9 May 2025.

**Results:** Three hundred and fifteen patients were included, of whom 104 met 2024 ACC/AHA guideline criteria for SRT eligibility at baseline. The SRT-eligible cohort was predominantly female (57%), with mean resting and Valsalva left ventricular outflow tract (LVOT) gradients of 63 ± 39 and 109 ± 42 mmHg, and all were in New York Heart Association (NYHA) class III. All baseline SRT-eligible patients became SRT-ineligible with aficamten therapy during study follow-up over a median of 42 days (IQR: 17, 49), except for one participant who withdrew from the study to pursue SRT (total of 3 participants withdrew). After dose titration, 3/104 (2.9%) remained guideline-eligible; by week 72 no patients met eligibility criteria. At maintenance, resting and Valsalva LVOT gradients improved by a least-squares mean of −41 mmHg ([95% CI −44 to −37]; *P<*0.0001) and −56 mmHg ([95% CI −62 to −51]; *P<*0.0001), respectively. Relative to baseline, NT-proBNP improved by 77% (95% CI 74 – 80%), high-sensitivity cardiac troponin I decreased by 38% (95% CI 30 – 46%), KCCQ-CSS improved by a mean of 20.2 (SD 19.3) points, and 95.2% of SRT-eligible patients had improved by ≥1 NYHA class. Overall, the safety profile was favorable, with 2 occurrences of left ventricular ejection fraction (LVEF) < 50% over 193.7 patient-years of follow-up (1 event per 100 patient-years), managed by down-titration. There were no baseline SRT-eligible patients who died or developed LVEF <40%.

**Conclusions:** Aficamten resolved guideline eligibility for SRT in nearly all baseline-eligible patients, with rapid and durable improvements in hemodynamics, symptoms, biomarkers and health status sustained for up to 3.5 years. Instances of LVEF <50% were rare and without clinical sequelae. These data support aficamten as a safe and effective alternative to SRT in oHCM.

**Registration:** **ClinicalTrials.gov, NCT04848506 (https://clinicaltrials.gov/study/NCT04848506); Date of registration: April 19, 2021**

**Clinical Perspective:** *What is new?:*

- Aficamten is a cardiac myosin inhibitor that improved symptoms and left ventricular outflow tract gradients in almost all patients who were eligible for septal reduction therapy (SRT) in the FOREST-HCM trial.
- Aficamten was safe with rare reduction in systolic function and no heart failure events.

*What are the clinical implications?:*

- Aficamten can be used as an alternative to patients who are eligible for SRT.

## Introduction

Hypertrophic cardiomyopathy (HCM) is the most common inherited cardiac disease, and left ventricular outflow tract (LVOT) obstruction is frequently present in symptomatic patients.^1^ Dynamic LVOT obstruction is independently associated with progression of heart failure symptoms, atrial fibrillation, stroke, and cardiovascular death, and relief of obstruction remains the principal therapeutic target in obstructive HCM (oHCM).^2^ Current international guidelines recommend septal reduction therapy (SRT), either surgical septal myectomy or alcohol septal ablation (ASA), for patients who remain in New York Heart Association (NYHA) functional class III or IV and have a resting or provocable LVOT gradient ≥50 mmHg despite maximally tolerated medical therapy.^3–5^

At dedicated, experienced, high-volume HCM centers, both septal myectomy and ASA are associated with in-hospital mortality rates of <1% and excellent long-term outcomes.^6–8^ However, outcomes in broader real-world practice vary substantially and remain dependent on procedural volume and institutional expertise. ^7–11^ Nevertheless, access to dedicated high-volume HCM centers remains limited globally, creating an unmet need for effective and broadly accessible alternatives to invasive septal reduction.^12^ Data from the SHaRe registry demonstrate that only a minority of patients with HCM ultimately undergo SRT, even at high-volume expert centers.^13^ ^14^

Cardiac myosin inhibitors, a new pharmacological class that attenuates myocardial hypercontractility by reducing actin–myosin cross-bridge formation, directly target the underlying sarcomeric pathophysiology of HCM.^15,16^ Mavacamten, the first-in-class agent, reduced guideline-based eligibility for SRT in the placebo-controlled VALOR-HCM trial at both 16 and 32 weeks.^17,18^ Aficamten is a next-in-class cardiac myosin inhibitor with a shorter plasma half-life and a broader therapeutic window, permitting more rapid up-titration and faster recovery from supratherapeutic exposure.^15,19^ In SEQUOIA-HCM, a pivotal phase 3 trial, aficamten produced significant improvements across the spectrum of oHCM disease burden, including complete hemodynamic response in 68% of patients, and a reduction in the amount of time spent eligible for SRT during the 24-week treatment period.^20,21^ ^22^ The long-term effect of aficamten in patients with oHCM and meeting guideline criteria for SRT has not been previously reported.

We hypothesized that chronic treatment with aficamten would reduce guideline-based eligibility for SRT in patients with oHCM, with a safety profile that compares favorably with the procedural risks of SRT. To test this hypothesis, we examined the SRT-eligible subgroup of the oHCM cohort in FOREST-HCM (Follow-Up Open-Label Research Evaluation of Sustained Treatment with Aficamten in Hypertrophic Cardiomyopathy; NCT04848506), the open-label long-term extension study of aficamten.

## METHODS

### Data availability

Qualified researchers may submit a request containing the research objectives, endpoints/outcomes of interest, statistical analysis plan, data requirements, publication plan, and qualifications of the researcher(s). In general, Cytokinetics, Incorporated, does not grant external requests for individual patient data for the purpose of reevaluating safety and efficacy issues already addressed in the product labeling. Requests are reviewed by a committee of internal advisors, and if not approved, may be further arbitrated by a Data Sharing Independent Review Panel. On approval, the information necessary to address the research question will be provided under the terms of a data sharing agreement. This may include anonymized individual patient data and/or available supporting documents, containing fragments of analysis code where provided in analysis specifications. Requests may be submitted to

### Design and Patients

FOREST-HCM is a multinational, open-label, long-term extension study evaluating the safety, tolerability and efficacy of aficamten in adults with HCM who completed a qualifying parent study (REDWOOD-HCM, SEQUOIA-HCM, MAPLE-HCM, or ACACIA-HCM). The study schema is presented in Supplemental Figure 1. The present analysis was restricted to patients with oHCM and follow-up through the maintenance dose visit (i.e., completed dose titration and achieved a stable maintenance dose) until 9 May 2025. Eligibility for SRT was prospectively defined according to the 2024 AHA/ACC guideline criteria ^3^ as the presence of a resting or provocable LVOT gradient ≥50 mmHg and NYHA functional class III or IV symptoms.

Patients meeting these criteria at baseline constituted the SRT-eligible cohort for the purposes of this analysis; the remainder formed the SRT-ineligible comparator group. The study was conducted in accordance with the Declaration of Helsinki and the International Council for Harmonisation Good Clinical Practice guidelines. The protocol was approved by the institutional review board or independent ethics committee at each participating site, and all participants provided written informed consent prior to any study-related procedures.

### Treatment

Aficamten was administered orally once daily, starting at 5 mg, with echocardiography-guided up-titration in 5-mg increments to a maximum dose of 20 mg based on site-read left ventricular ejection fraction (LVEF) and Valsalva LVOT gradient. Dose reductions were mandated for LVEF <50% but ≥40%, and treatment interruption for LVEF <40%.

### Clinical Outcomes

The primary outcome was the proportion of baseline SRT-eligible patients who remained eligible at each study visit. Secondary outcomes included time to first SRT-ineligibility, changes from baseline in resting and Valsalva LVOT gradients, LVEF, N-terminal pro-B-type natriuretic peptide (NT-proBNP), high-sensitivity cardiac troponin I (hs-cTnI), Kansas City Cardiomyopathy Questionnaire Clinical Summary Score (KCCQ-CSS), and NYHA functional class. The principal safety outcome was the exposure-adjusted incidence rate of LVEF <50% by site-read echocardiography, with LVEF <40% as a secondary safety threshold.

### Statistical Analysis

Continuous variables were summarized as mean (SD) or median (IQR), as appropriate, and categorical variables as n (%). Change from baseline in continuous variables were modeled using a mixed model for repeated measures (MMRM), with baseline value, visit, treatment subgroup (SRT-eligible vs. SRT-ineligible), treatment subgroup-by-baseline value interaction and treatment subgroup-by-visit interaction as fixed effects. Biomarker data (NT-proBNP and troponin) were log-transformed and reported as geometric means and ratios with 95% CIs.

Exposure-adjusted incidence rates (EAIR) for LVEF <50% were calculated as the number of patients with a first event divided by total patient-years of exposure and reported per 100 patient-years. Two-sided P-values <0.05 were considered nominally statistically significant. Analyses were performed using SAS Enterprise Guide version 8.3.

Dr Masri had full access to all the data in the study, verified the underlying data, and takes responsibility for the integrity of the data and the accuracy of the data analysis.

## RESULTS

### Baseline Characteristics

A total of 315 patients with oHCM were included in the analysis. Of these, 104 patients (33%) met guideline criteria for SRT at baseline (SRT-eligible), while 211 were SRT-ineligible at baseline (NYHA I/II, or LVOT gradient below threshold). Baseline characteristics are presented in Table 1. Compared with the SRT-ineligible cohort, the SRT-eligible cohort had a higher body mass index (30.0 ± 4.6 vs. 28.3 ± 3.8 kg/m², *P=*0.001) and was more likely to be female (56.7% vs. 38.9%, *P=*0.003). All SRT-eligible patients were in NYHA class III at baseline, compared with 8.1% of SRT-ineligible patients (*P<*0.001). SRT-eligible patients had higher mean resting (63 ± 39 vs. 52 ± 35 mmHg) and Valsalva (109 ± 42 vs. 86 ± 40 mmHg) LVOT gradients (*P=*0.019 and p<0.001, respectively). Background calcium channel blocker therapy was more common among SRT-eligible participants (31.7% vs. 13.7%, *P<*0.001).

**Table 1.** Demographics and Baseline Characteristics.

|  | Overall (n=315) | SRT-eligible<br>(n=104) | SRT-ineligible<br>(n=211) | *P*-value |
| --- | --- | --- | --- | --- |
| Age, y, mean (SD) | 60.7 (12.7) | 62.1 (12.7) | 60.0 (12.7) | 0.175 |
| Age, y, median (Q1, Q3) | 62.0 (53.0, 70.0) | 65.0 (54.0, 72.0) | 61.0 (53.0, 69.0) | 0.105 |
| Female sex, n (%) | 141 (44.8) | 59 (56.7) | 82 (38.9) | 0.003 |
| White race, n (%) | 296 (94.0) | 96 (92.3) | 200 (94.8) | 0.444 |
| BMI, kg/m<sup>2</sup>, mean (SD) | 28.8 (4.1) | 30.0 (4.6) | 28.3 (3.8) | 0.001 |
| Positive family history of HCM, n (%) | 82 (26.0) | 20 (19.2) | 62 (29.4) | 0.054 |
| NYHA functional class, n (%) |  |  |  | <0.0001 |
| Class I | 11 (3.5) | 0 | 11 (5.2) |  |
| Class II | 183 (58.1) | 0 | 183 (86.7) |  |
| Class III | 121 (38.4) | 104 (100.0) | 17 (8.1) |  |
| Class IV | 0 | 0 | 0 |  |
| Background HCM therapy, n (%) |  |  |  |  |
| Beta-blocker monotherapy | 198 (62.9) | 56 (53.8) | 142 (67.3) | 0.020 |
| CCB monotherapy (direct cardiac) | 62 (19.7) | 33 (31.7) | 29 (13.7) | 0.0002 |
| Disopyramide monotherapy | 44 (14.0) | 16 (15.4) | 28 (13.3) | 0.611 |
| Two or more background medications | 49 (15.6) | 16 (15.4) | 33 (15.6) | 0.953 |
| No background HCM medication | 62 (19.7) | 17 (16.3) | 45 (21.3) | 0.296 |
| LVEF, %, mean (SD) | 68.4 (5.8) | 69.5 (5.5) | 67.9 (5.9) | 0.022 |
| Resting LVOT gradient, mmHg, mean (SD) | 55.8 (36.9) | 62.7 (39.3) | 52.3 (35.2) | 0.019 |
| Valsalva LVOT gradient, mmHg, mean (SD) | 93.7 (42.2) | 109.1 (41.9) | 86.1 (40.3) | <0.0001 |
| NT-proBNP, pg/mL, median (Q1, Q3) | 778 (346, 1529) | 801 (380, 1573) | 772 (332, 1529) | 0.527 |
| hs-Troponin I, ng/L, median (Q1, Q3) | 11.0 (5.8, 19.6) | 11.1 (5.4, 26.8) | 10.6 (5.9, 19.1) | 0.182 |
| Maintenance-dose, n (%) |  |  |  | 0.027 |
| 5 mg | 20 (6.3) | 1 (1) | 19 (9.0) |  |
| 10 mg | 61 (19.4) | 18 (17.3) | 43 (20.4) |  |
| 15 mg | 104 (33.0) | 35 (33.7) | 69 (32.7) |  |
| 20 mg | 130 (41.3) | 50 (48.1) | 80 (37.9) |  |

All echo measurements were from site-reads.
Modified Efficacy Analysis Set and Safety Analysis Set are presented. SRT eligibility defined by 2020 AHA/ACC guidelines
(NYHA III/IV and resting or provoked LVOT gradient $\geq 50$ mmHg).
BMI indicates body mass index, CCB, calcium channel blocker; HCM, hypertrophic cardiomyopathy; hs, high-sensitivity;
LVEF, left ventricular ejection fraction; LVOT, left ventricular outflow tract; NT-proBNP, N-terminal pro-B-type natriuretic
peptide; NYHA, New York Heart Association; Q1, 25th percentile; Q3, 75th percentile; SD, standard deviation; SRT, septal
reduction therapy.

### Resolution of SRT Eligibility

The proportion of baseline SRT-eligible patients who continued to meet guideline criteria for SRT declined rapidly and progressively during aficamten titration and remained low throughout long-term follow-up (Figure 1). By the first titration visit, 76 of 104 evaluable SRT-eligible patients (73.1%) remained eligible; by the second titration visit, 40 (38.5%); and by the third titration visit, 14 (13.5%). At the first maintenance visit, 3 of 104 patients (2.9%) remained eligible for SRT. By week 72, no baseline SRT-eligible patient met guideline criteria for SRT, with similar findings through week 168. Interpretation of later time points is limited by the smaller number of patients with available follow-up data; only 1 patient (1/45; 2.2%) transiently met SRT eligibility criteria at week 96 before again becoming ineligible. The median time to first SRT-ineligibility was 42 days (IQR 17–49 days; range, 14–261). At the time of first SRT-ineligibility, 58% of patients became ineligible due to LVOT gradient reduction, 5% due to NYHA class improvement, and 38% because of both LVOT gradient reduction and NYHA class improvement. Three baseline SRT-eligible patients withdrew from the study at weeks 38, 47, and 56. The patient who withdrew at week 56 opted to pursue SRT rather than remain in the study.

**Figure 1:**
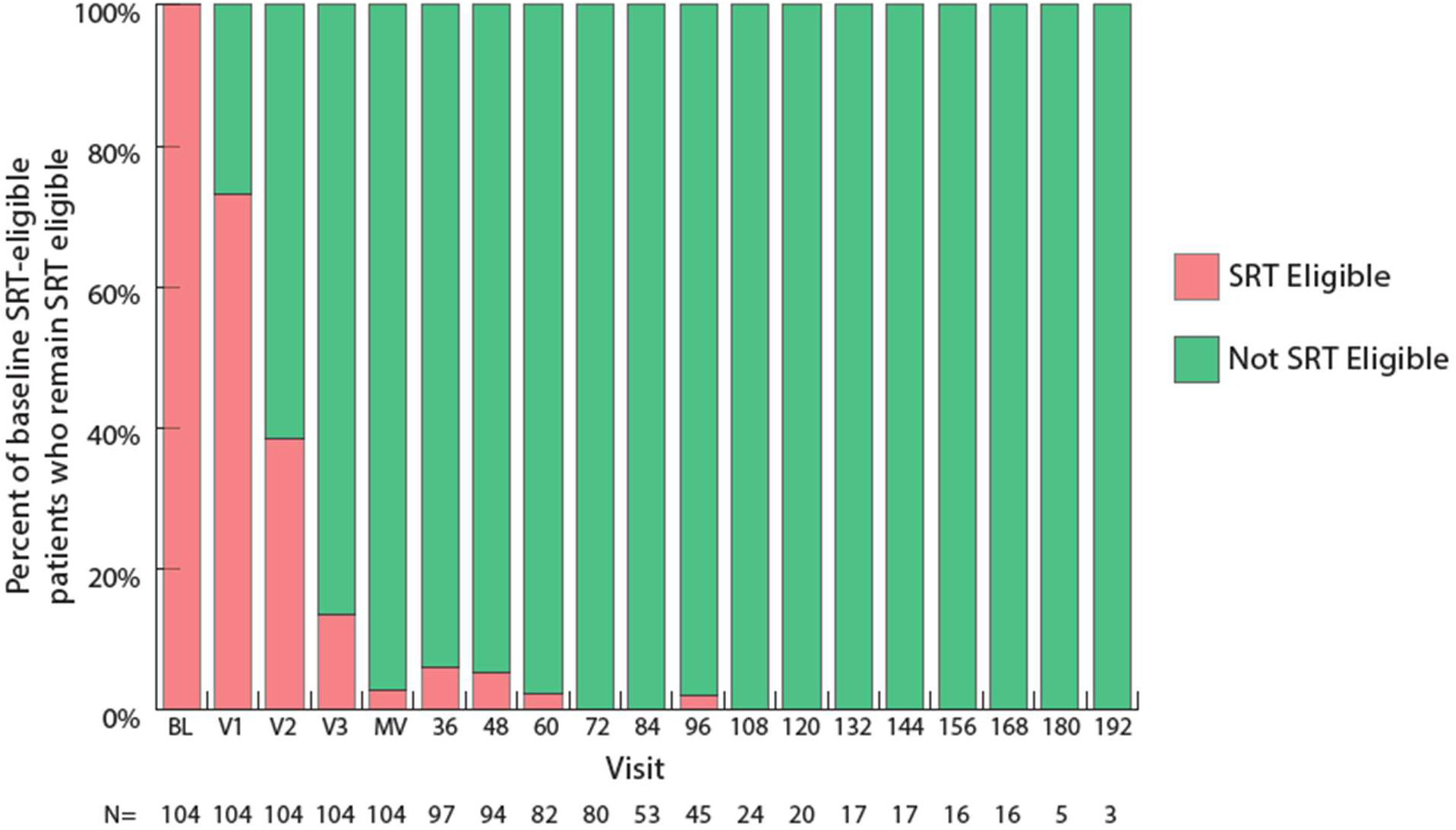
Proportion of baseline SRT-eligible patients who remained SRT-eligible during aficamten treatment in FOREST-HCM. SRT eligibility was defined per 2020 AHA/ACC guidelines (NYHA III/IV and resting or provoked LVOT gradient ≥50 mmHg). All 104 evaluable patients (100%) were SRT-eligible at baseline; only 2.9% remained eligible at the maintenance visit, and no patient met eligibility criteria from week 72 onwards (with a single transient exception at week 96).AHA/ACC indicates American Heart Association/American College of Cardiology, LVOT left ventricular outflow tract, NYHA New York Heart Association, SRT septal reduction therapy.

The other 2 patients withdrew because of perceived lack of efficacy (week 38) and ischemic colitis (week 47); both were SRT-eligible at withdrawal and were lost to follow-up. In the baseline SRT-ineligible group, only 7 patients became transiently eligible for SRT during treatment, of whom 1 was eligible for SRT at the last follow-up visit.

### Hemodynamic Response

Aficamten produced large, sustained reductions in LVOT obstruction in both cohorts (Figure 2). In the SRT-eligible group, the resting LVOT gradient decreased from a baseline mean of 63 mmHg (SD 39) to 16 mmHg (SD 14) at the maintenance visit (LSM change, −40.7 mmHg, [95% CI, −44 to −37]; *P<*0.0001). Valsalva-provoked gradient decreased from 109 mmHg (SD 42) to 40 mmHg (SD 28) (LSM change, −56.2 mmHg, [95% CI, −62 to −51]; *P<*0.0001). Similar reductions were observed in the SRT-ineligible cohort (resting LSM change −39 mmHg; Valsalva −55 mmHg). Reductions in LVOT gradients were detected by the first titration visit, increased through the end of titration and were maintained throughout the study period (up to 168 weeks of follow-up).

**Figure 2.**
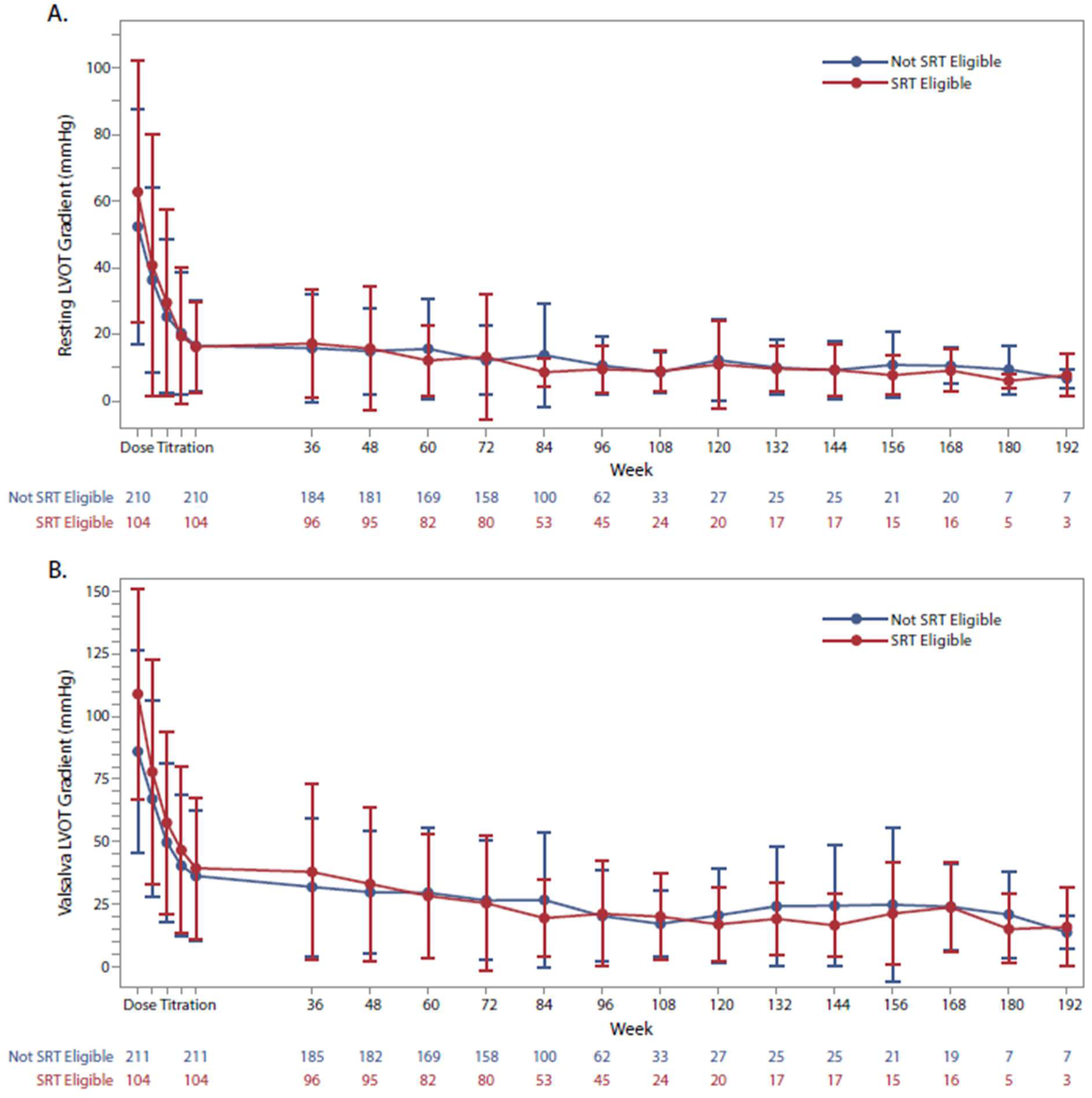
Resting and Valsalva LVOT Gradients over Time in SRT-Eligible and SRT-Ineligible Patients. (A) Resting left ventricular outflow tract gradient and (B) Valsalva left ventricular outflow tract gradient. LVOT indicates left ventricular outflow tract.

### Symptoms and Health Status

Symptom burden and patient-reported health status improved rapidly and durably on aficamten (Figure 3). In the SRT-eligible cohort, KCCQ-CSS increased from a baseline mean of 58.0 ± 19.1 to 78.2 ± 18.1 at the maintenance visit (change: 20.2 ± 19.3 points), with additional modest improvement during longer-term follow-up. A ≥1-class improvement in NYHA functional class was achieved in 89.2% of SRT-eligible patients by the second titration visit, 95.2% at the maintenance visit, and 100% of evaluable patients from week 108 onward. At the maintenance visit, 34 of 104 SRT-eligible patients (32.7%) had improved to NYHA class I, 65 (62.5%) to class II, and 5 (4.8%) remained in class III; no patients were in class IV.

**Figure 3:**
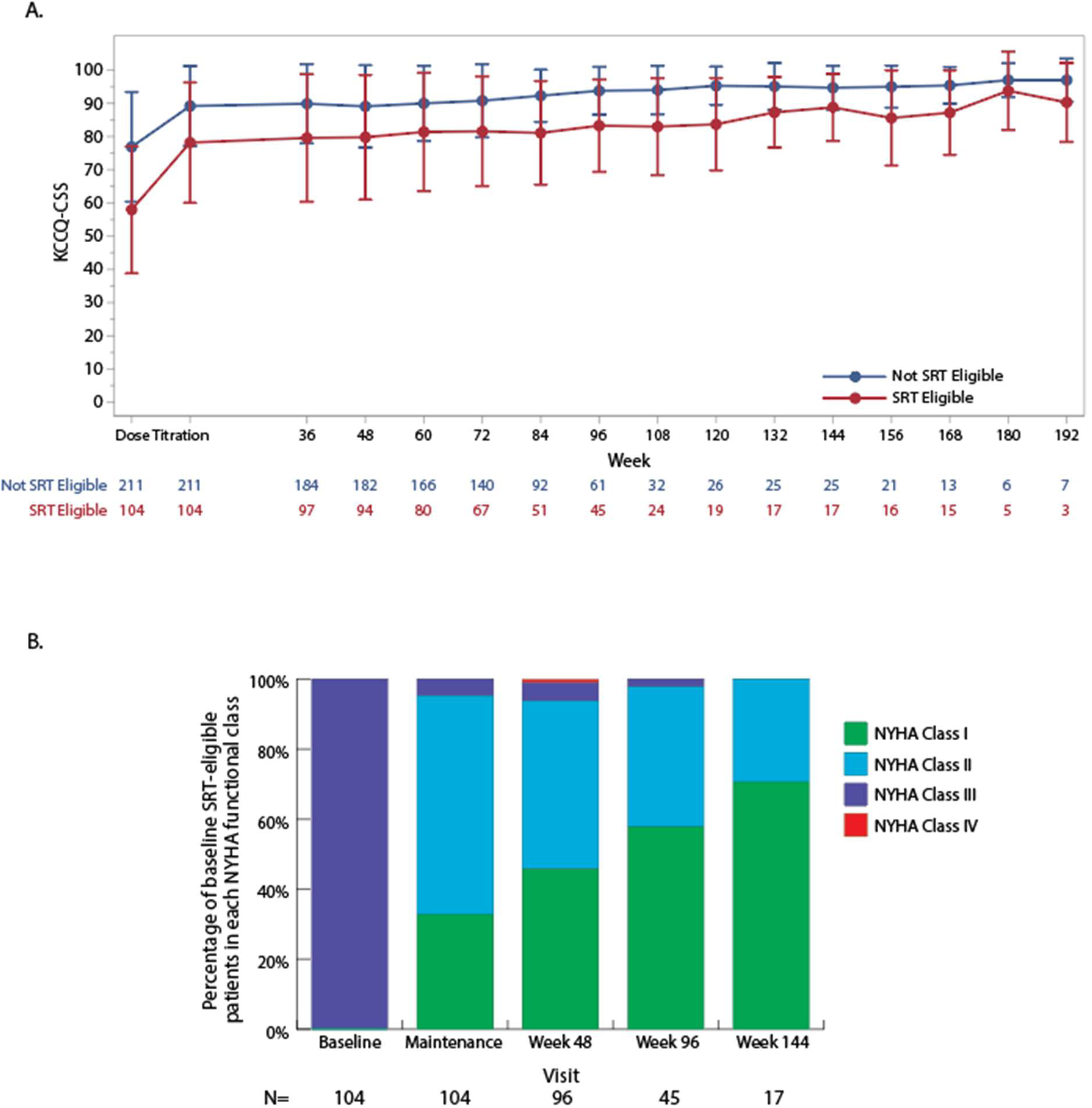
Health status and symptomatic response over time (change in KCCQ-CSS and NYHA status). KCCQ-CSS indicates Kansas City Cardiomyopathy Questionnaire Clinical Summary Score, NYHA New York Hear Association.

### Cardiac Biomarkers

Serum concentration of cardiac biomarkers decreased rapidly and to a similar extent in both cohorts. In the SRT-eligible group, NT-proBNP declined from a baseline geometric mean of 725 pg/mL to 165 pg/mL at the maintenance visit, representing a 77% ([95% CI 74% - 80%]; *P<*0.0001) relative reduction. The hs-cTnI decreased from 14.7 ng/L to 9.0 ng/L, corresponding to a 38% (95% CI 30% - 46%) relative reduction. Both effects were sustained throughout long-term follow-up.

#### Safety

During a total of 193.7 patient-years of follow-up in the SRT-eligible cohort, 2 patients had an LVEF <50% (exposure-adjusted incidence rate, 1.03 per 100 patient-years). In the SRT-ineligible cohort, 10 patients had an LVEF <50% over 348.4 patient-years (2.87 per 100 patient-years), resulting in an overall occurrence of 12 of 317 patients (3.8%) over 542.0 patient-years (2.2 per 100 patient-years) in the oHCM cohort. No patient in either cohort had an LVEF <40% at any time during follow-up. The cross-sectional prevalence of LVEF <50% at any post-baseline visit remained low, never exceeding 1.9% in the SRT-eligible cohort, and declined to 0% by week 36, and remained at 0% through week 192. LVEF excursions were managed according to protocol through dose reduction or temporary interruption, and no patient required permanent discontinuation of aficamten for reduced LVEF.

## DISCUSSION

In this analysis of the oHCM cohort of FOREST-HCM, the long-term extension study of aficamten, we make 3 principal observations. First, among patients who met guideline criteria for SRT at baseline, long-term aficamten therapy rapidly resolved guideline eligibility in nearly all evaluable patients. Second, improvements in LVOT obstruction, symptoms, patient-reported health status, and cardiac biomarkers were rapid, substantial, and sustained through more than 3 years of follow-up. Third, safety with respect to LVEF was excellent: only 2 LVEF <50% events occurred during 193.7 patient-years of SRT-eligible exposure (exposure-adjusted event rate, approximately 1 per 100 patient-years), were without clinical consequence, and no LVEF <40% events occurred. Importantly, the observed safety profile compares favorably with contemporary benchmarks for in-hospital event rates and morbidity after SRT in real-world U.S. practice.^7,9^ ^8,237,9^Symptomatic improvement through relief of LVOT obstruction has been the cornerstone of oHCM management for more than 60 years. Since the initial Morrow myectomy, outcomes at dedicated high-volume centers have steadily improved. At the most experienced programs, in-hospital mortality after isolated myectomy has been reported to be as low as 0.4%, and long-term survival may be indistinguishable from that of the age-matched general population.^8,23^ These benchmarks represent the ideal and are appropriately cited in guideline recommendations for SRT. However, such outcomes are not achieved universally, as they depend heavily on operator experience and are not scalable to the extent required for all, or even most, patients with oHCM. Altibi and colleagues analyzed 12,065 septal myectomy admissions between 2010 and 2019 in a nationally representative U.S. sample and reported an all-comers in-hospital mortality of 4.0%. Two-thirds of hospitals performed 5 or fewer myectomies per year; 10% of hospitals had a myectomy mortality ≥10%; and 3.7% had a mortality rate of 100%. Even among hospitals in the highest-volume decile, in-hospital adverse events, including stroke, acute kidney injury, complete heart block, pacemaker implantation, mechanical circulatory support, and readmission within 30-days, occurred in more than 10% of patients.^7,9^

An earlier analysis of the 2003–2011 U.S. Inpatient Sample reported an even higher overall mortality (5.9%) with substantial heterogeneity across centers.^7^ ASA appears to confer significantly lower overall mortality even in real-world practice (approximately 1.2%) but is associated with higher long-term rates of pacemaker implantation (11–12% in most series) and, in some studies, a lower likelihood of complete hemodynamic response. ^7,9,10^ In the context of these real-world data, the benchmark for a medical alternative to SRT is the achievement of hemodynamic and symptomatic benefit while avoiding the procedural morbidity and mortality that cannot be reliably guaranteed outside a limited number of tertiary centers.

The current data suggest that aficamten satisfies this standard. In FOREST-HCM, nearly all patients who were SRT-eligible at baseline achieved SRT-ineligibility, most within 6 weeks of initiating therapy. This efficacy is consistent with, and extends follow-up of, the effects observed in the SEQUOIA-HCM responder analysis, in which 88% of SRT-eligible patients became SRT-ineligibile, at 24 weeks,^21^ and is broadly similar to the approximately 80% conversion rate observed with mavacamten in VALOR-HCM at 16 weeks.^18^ Importantly, the durability of these effects has not previously been demonstrated for a cardiac myosin inhibitor in a purely SRT-eligible population. At every scheduled assessment from week 72 through week 168, only one patient who was SRT-eligible at baseline transiently met guideline criteria for SRT. These findings are also consistent with data from a FOREST-HCM standard-of-care withdrawal substudy, in which withdrawal of background medications was safely achieved in most patients receiving aficamten without loss of hemodynamic benefits.^24^

The safety profile is particularly relevant to the hypothesis that aficamten may represent a safer alternative to SRT for many patients. In FOREST-HCM, the event rate of LVEF <50% among SRT-eligible patients was 1.03 per 100 patient-years; in the overall oHCM population, the rate was 2.21 per 100 patient-years. No patient had LVEF <40% at any time during follow-up, required permanent discontinuation of aficamten for reduced LVEF, or experienced clinical decompensation associated with a transient reduction in LVEF. By comparison, Altibi et al report an overall in-hospital mortality rate of 4.0% after SRT (a one-time event occurring around the time of the procedure), rising to 5.7% in low-volume centers.^9^ In-hospital adverse events, including stroke, renal failure requiring dialysis, ventricular septal defect, and permanent pacemaker implantation, occurred in 21.3% of patients at low-volume centers and 12.6% at high-volume centers; 30-day readmission rates ranged from 9.7% to 17.1%.^9^ The periprocedural risk of SRT is front-loaded and concentrated during the first 30 days, whereas the risks of chronic aficamten treatment are mild, manageable through dose titration, and reversible in all cases.

Although direct comparisons between FOREST-HCM and real-world reports are limited, patients in FOREST-HCM were managed by their treating physicians, who were able to integrate clinical data into dosing and titration decisions without reliance on an echocardiographic core-laboratory. As such, FOREST-HCM approximates routine clinical practice, and the observed safety profile supports the contention that, for an individual patient referred for SRT in a community setting, initiation of aficamten may carry a lower absolute short-term risk than proceeding to invasive septal reduction outside a high-volume center.

Our findings also have implications for equity of access. Current guidelines emphasize referral of SRT candidates to dedicated high-volume HCM centers, but such centers are few and unevenly distributed. Patients who cannot travel, have prohibitive perioperative risk, or prefer to avoid invasive therapy currently have limited options. The reliable and durable SRT avoidance demonstrated here, coupled with a favorable safety profile achievable with echocardiographic dose titration that can be performed at most secondary centers, suggests that aficamten may materially expand access to effective, guideline-concordant care for oHCM. At the same time, long-term pharmacologic therapy introduces important tradeoffs, including ongoing treatment costs, the need for continued echocardiographic monitoring and follow-up, and lifelong dependence on medical therapy compared with the potential durability of a one-time septal reduction procedure, which may remain preferable for some patients.

### Limitations

Several limitations should be acknowledged. FOREST-HCM is a single-arm, open-label extension study; there is no concurrent randomized control, and comparisons with historical SRT outcomes must be interpreted cautiously given differences in patient populations, treatment era, and ascertainment. Patients enrolled in FOREST-HCM had completed a qualifying parent trial and may therefore represent a selected population with demonstrated tolerance of aficamten. The SRT-eligible cohort comprised patients who either chose not to undergo SRT before enrollment or did not have access to SRT and may therefore differ systematically from all-comer SRT candidates. Finally, longer-term follow-up is required to determine whether the hemodynamic and structural benefits observed with cardiac myosin inhibition translate into improvements in hard cardiovascular endpoints, including sudden death, stroke, and heart failure hospitalization.

## CONCLUSIONS

In the oHCM cohort of FOREST-HCM, long-term aficamten treatment resolved guideline eligibility for SRT in nearly all patients who were SRT-eligible at baseline and produced rapid, substantial, and durable improvements in hemodynamics, symptoms, health status and cardiac biomarkers sustained through more than 3 years of follow-up. The occurrence of LVEF <50% was rare, at an EAIR of 1.03 per 100 patient-years in the SRT-eligible cohort, there were no LVEF <40% events, and the results compare favorably with contemporary real-world in-hospital mortality and morbidity after SRT. These data support aficamten as an effective and safe alternative to invasive septal reduction in patients with oHCM over the intermediate term.

## Acknowledgments

The study team and the authors thank all patients who participated in this study and their families. They also thank all study sites and research personnel for their contributions. Editorial support was provided by David Sunter, PhD, on behalf of Engage Scientific Solutions, and was funded by Cytokinetics, Incorporated.

## Supplemental Material

Figures S1–S2

## Non-standard Abbreviations and Acronyms

ASA: Alcohol septal ablation
CNIC: Centro Nacional de Investigaciones Cardiovasculares
EAIR: Exposure-adjusted incidence rates
IRCCS: Istituto di Ricovero e Cura a Carattere Scientifico
KCCQ-CSS: Kansas City Cardiomyopathy Questionnaire Clinical Summary Score
LSM: Least squares mean
LVEF: Left ventricular ejection fraction
LVOT: Left ventricular outflow tract
MMRM: Mixed model for repeated measures
NYHA: New York Heart Association
SRT: Septal reduction therapy

